# Discernment of Mediator and Outcome Measurement in the PACE trial

**DOI:** 10.1101/2021.01.25.21250436

**Authors:** Ewan Carr, Silia Vitoratou, Trudie Chalder, Kimberley Goldsmith

**Affiliations:** Department of Biostatistics and Health Informatics, Institute of Psychiatry, Psychology and Neuroscience, King’s College London, London, United Kingdom; Academic Department of Psychological Medicine, Institute of Psychiatry, Psychology and Neuroscience, Weston Education Centre, King’s College London, London, United Kingdom; South London and Maudsley NHS Foundation Trust, London, United Kingdom

**Keywords:** Mediation, confirmatory factor analysis, construct overlap, latent trait

## Abstract

**Background:** When measuring latent traits, such as those used in psychology and psychiatry, it can be unclear whether the instruments used are measuring different concepts. This issue is particularly important in context of mediation analysis, since for a sound mediation hypothesis the mediator and outcome should be distinct. We sought to assess the extent of measurement overlap between mediators and outcomes in the PACE trial (n=640).

**Methods:** Potential measurement overlap was assessed using generalised linear latent variable models where confirmatory factor models quantified the extent to which the addition of cross-loading items resulted in significant improvements in model fit.

**Results:** Out of 26 mediator-outcome pairs considered, only six showed evidence of cross-loading items, supporting the suggestion that mediator and outcome constructs in the PACE trial were conceptually distinct.

**Conclusions:** This study highlights the importance of assessing measurement overlap in mediation analyses with latent traits to ensure mediator and outcome instruments are distinct.

## Background

Studies in psychiatry and psychology are often interested in unobserved constructs such as behaviour or cognition that are captured using multiple items from a questionnaire. These constructs can be measured within the generalised linear latent variable models framework (Bartholomew et al., 2011; Skrondal & Rabe-Hesketh, 2004), consisting of two parts: the measurement part and the structural part of the model. The measurement part refers to the observed items which designate (‘load’ onto) a latent variable (the estimated magnitudes of these relationships are referred to as the ‘factor loadings’); the structural part refers to the latent variables and their relationships.

The measurement part of the model is typically specified based on theory or prior empirical research. The assignment of observed items to latent constructs may be straightforward if, for example, the items are drawn from an established psychological scale, or there is some other strong theoretical motivation for including, on a given instrument, some items over others. However, this is not always the case and there may be some ambiguity about which items to include on a given instrument. A single observed item might be related to two or more latent constructs, represented in the model by two or more statistically significant factor loadings. This arrangement would produce ‘cross-loadings’, where a single item loads initially onto the intended latent construct (the ‘primary loading’) as well as onto a second latent construct (the ‘cross-loading’).

In this study we are concerned with what happens when two measurement instruments share several cross-loading items and how our interpretations should be adjusted accordingly. While overlap may exist in the structural part of the model (e.g. Schaufeli & Bakker, 2004), our focus here is on overlap in the measurement part of the model: do the items measuring one construct also serve as indicators of another? We are particularly interested in how measurement overlap affects the interpretation of mediational hypotheses. Mediation is where a ‘third variable’ transmits the effect of one variable to another and is thus “intermediate in the causal sequence relating an independent variable to a dependent variable” (p. 1, MacKinnon, 2008). The single mediator model in a clinical trial setting postulates that the effect of a randomly-assigned treatment (*R*) on a continuous outcome (*Y*) is mediated by a third variable (*M*) (MacKinnon, 2008). Since Baron and Kenny’s widely-cited paper describing mediation and moderation in psychology (1986), methods for mediation analysis have grown in sophistication (Emsley et al., 2010; K. Goldsmith et al., 2018; K. A. Goldsmith et al., 2018; Preacher, 2015; VanderWeele et al., 2014, 2012). It is now recognised that single mediator model makes several inferential assumptions (MacKinnon, 2008) including temporal precedence (*R* occurs before *M* which occurs before *Y*), that the temporal ordering is appropriate for the hypothesised mediational chain, and that there are no omitted influences (i.e. other variables influencing *R, M*, or *Y*, either observed or unobserved). Here we consider the additional (and often untested) assumption that *M* and *Y* represent distinct theoretical constructs. If the mediator represents an earlier assessment of the outcome then the model interpretation changes; the mediator no longer represents a ‘third variable’ in the mediational chain.

Psychological studies often involve latent constructs that necessarily share common themes and have the potential to overlap. Kaufman et al. (2005), for example, considered potential mediators of CBT for adolescents with depressive symptoms. Their potential mediators included the Automatic Thoughts Questionnaire (ATQ) (Hollon & Kendall, 1980) which shares similar themes and question wordings as their outcome measure, the Beck Depression Inventory (BDI-II) (Beck et al., 1996). In particular, regarding notions of worthlessness, self-dislike, and self-criticalness. This raises the question of whether these two constructs are truly distinct. Identifying potential overlap is not always straightforward. Given the difficulties of distinguishing and measuring some latent traits, cross-loading may occur even among items that do not appear, on face value, to be conceptually similar. As we describe below, many of the mediators and outcomes studied in this paper do not share common items or wordings but nonetheless could be seen to share similar themes.

This paper was motivated by an existing mediation analysis that tested whether unhelpful cognitions and behaviours mediated the effect of cognitive behavioural therapy (CBT) in treating patients with chronic fatigue syndrome (Chalder et al., 2015). Drawing on data from the PACE trial (White et al., 2011), this analysis considered whether the effect of treatment allocation (*R*; 0 weeks) on fatigue (*Y*; 52 weeks) was mediated by variables measured at 12 weeks using the Cognitive Behavioural Responses Questionnaire (CBRQ; Ryan et al. (2017)). For example, illustrated in Figure 1, 34% of the effect of CBT on fatigue was transmitted via ‘fear avoidance’ beliefs.

**Figure 1.**
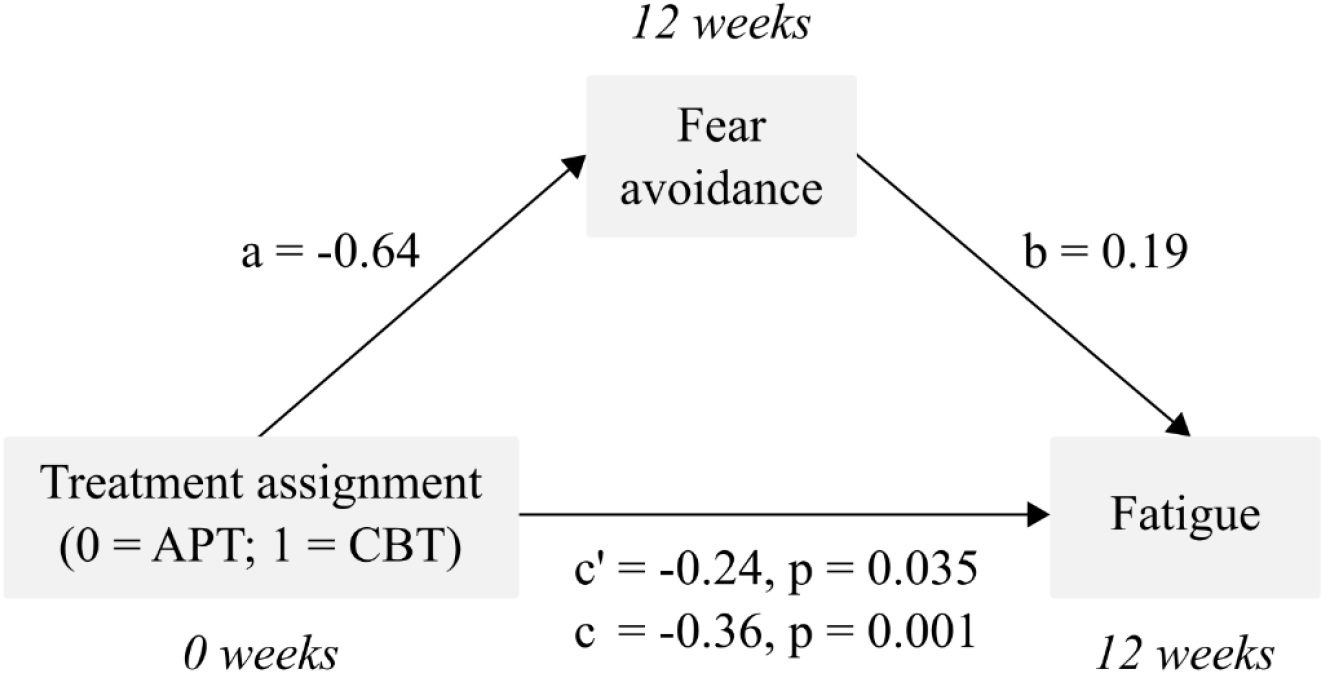
Standardised effects in mediation models, reproduced from Chalderet et al. 2015 (p. 150). Notes. CBT = cognitive behaviour therapy; APT = adaptive pacing therapy. Besides treatment, models included: centre, Standardised Clinical Interview for DSM-TV (SCID) depression status, London criteria for myalgic encephalomyelitis status, International Chronic Fatigue Syndrome (CFS) criteria, baseline measures of both outcome variables, baseline Work and Social Adjustment Scale, SCID anxiety disorder status, age, sex, CFS group membership, receipt of benefits, benefits in dispute, physical illnessattribution, fibromyalgia status, illness duration, Jenkins Sleep Score, employment status, body-mass index, and physical symptoms (PHQ-15) score.

The mediators and outcomes used in the PACE trial were purposefully chosen to represent distinct theoretical constructs. The questionnaires used to measure these constructs had previously been psychometrically evaluated and found to be reliable and valid (Cella et al., 2011; Chalder et al., 1993; McHorney et al., 1993; Ryan et al., 2017). This first step is critical: careful selection of appropriate constructs can help avoid ambiguity about which items correspond to which constructs and minimise potential overlap at the outset. However, despite these efforts, there remains the possibility of overlap between mediator and outcome constructs and the possibility that the identified mechanisms could in part be explained by measurement overlap (i.e. the mediator and outcome ‘measuring the same thing’). For example, items measuring the mediators include “I stay in bed to control my symptoms” and “When I experience symptoms, I rest” while items measuring the fatigue outcome include “Do you need to rest more?” and “Do you have problems with tiredness?”

This paper presents a procedure to assess the extent of measurement overlap between the mediators and outcomes used in the PACE trial to address the question: were the chosen mediators and outcomes ‘measuring the same thing?’ To our knowledge, no previous papers have addressed measurement overlap in the context of mediation analysis. We expect to find minimal overlap since the included instruments were chosen to represent distinct theoretical constructs and have previously been shown to be psychometrically reliable and valid. We note that our aim is to assess measurement overlap and not to re-evaluate the structure of these constructs, which has been addressed in previous studies (Cella et al., 2011; Chalder et al., 1993; McHorney et al., 1993; Ryan et al., 2017).

## Methods

### Data

The PACE trial was a four-arm randomised trial designed to assess the effectiveness of rehabilitative treatments for chronic fatigue syndrome (White et al., 2011). 640 patients were recruited from six specialist chronic fatigue syndrome clinics in the UK National Health Service between March 2005 and November 2008. Eligibility criteria, trial methods and results are described elsewhere (White et al., 2011). For this analysis we used mediator data gathered at 12 weeks post-randomisation and outcome data gathered at 52 weeks post-randomisation, consistent with the published mediation analysis of PACE (Chalder et al., 2015).

### Measures

We considered seven mediating factors consistent with those included in the PACE mediation analysis (Chalder et al., 2015), derived from questions on the Cognitive Behavioural Responses Questionnaire (Ryan et al., 2017). The five *cognitive* mediators were ‘fear avoidance,’ ‘catastrophising,’ ‘damage,’ ‘embarrassment avoidance,’ and ‘symptom focusing.’ Two *behavioural* mediators were ‘all-or-nothing behaviour’ and ‘behavioural avoidance.’ For this analysis, we included both the original versions of these factors as well as shortened versions (3-item) described in Ryan et al. (2017), with the exception of ‘catastrophising’ for which a short version was not available.

Two outcomes were considered, following the original PACE mediation analysis, both of which have previously been psychometrically validated. Fatigue was measured by the 11-item Chalder Fatigue Questionnaire (CFQ) (Cella et al., 2011; Chalder et al., 1993). Physical function was measured by the physical function subscale (PF, 10 items) of the Medical Outcomes Study Short Form Health Survey (SF-36) (McHorney et al., 1993). The factor structure and question wordings for each mediator and outcome are presented in Supplementary Table 1.

### Statistical analyses

We assessed overlap between the mediators and outcomes by fitting three sets of confirmatory factor models, described below. There were 13 mediating factors (7 factors plus 6 short versions) and two outcomes giving a total of 26 mediator-outcome pairs. Overall model fit was assessed based on the comparative fit index (CFI, with preferred values higher than 0.90) (Bentler, 1990), the root mean square error of approximation (RMSEA, with preferred values less than 0.08) (Browne & Cudeck, 1993), and the relative χ^2^ (ratio of χ^2^ to model degrees of freedom). Close fit was indicated by CFI ≥ 0.95, RMSEA ≤ 0.05 and relative χ^2^ close to 2 (Kaplan, 2008).

Changes in model fit – for example, after adding a cross-loading to a model – were assessed using the DIFFTEST procedure in Mplus (Asparouhov & Muthén, 2006). This is a procedure for testing nested models with mean and variance adjusted χ ^2^ statistics. In our example, this involved fitting two models: the unrestricted model (*H*_*1*_) where the cross-loading was freely estimated; and the restricted model (*H*_*0*_) where the cross-loading parameter was constrained to 0. All models were fitted in Mplus 7.3 (Muthén & Muthén, 2015) using R version 4.0.3 (R Core Team, 2017) and the MplusAutomation package (Hallquist & Wiley, 2018). All models were estimated using the robust weighted least squares (WLSMV) estimator.

Before investigating overlap between constructs, we made some assumptions about the individual factors. Firstly, as these factors had previously been psychometrically evaluated, we assumed that each factor would independently achieve good overall model fit. Secondly, since our aim was to assess construct overlap, not the structure of individual scales, we assumed the psychometric structure of each factor was known and appropriate at the outset.

In Step 1, we estimated a single factor confirmatory factor model (uni-dimensional CFA) for each mediator-outcome pair (Figure 2, Step 1). This was a model where a single continuous latent variable was measured by the observed items for *both* the mediator and outcome. For this model we hypothesised that overall model fit would be poor since these latent variables were expected to be measuring distinct constructs.

**Figure 2.**
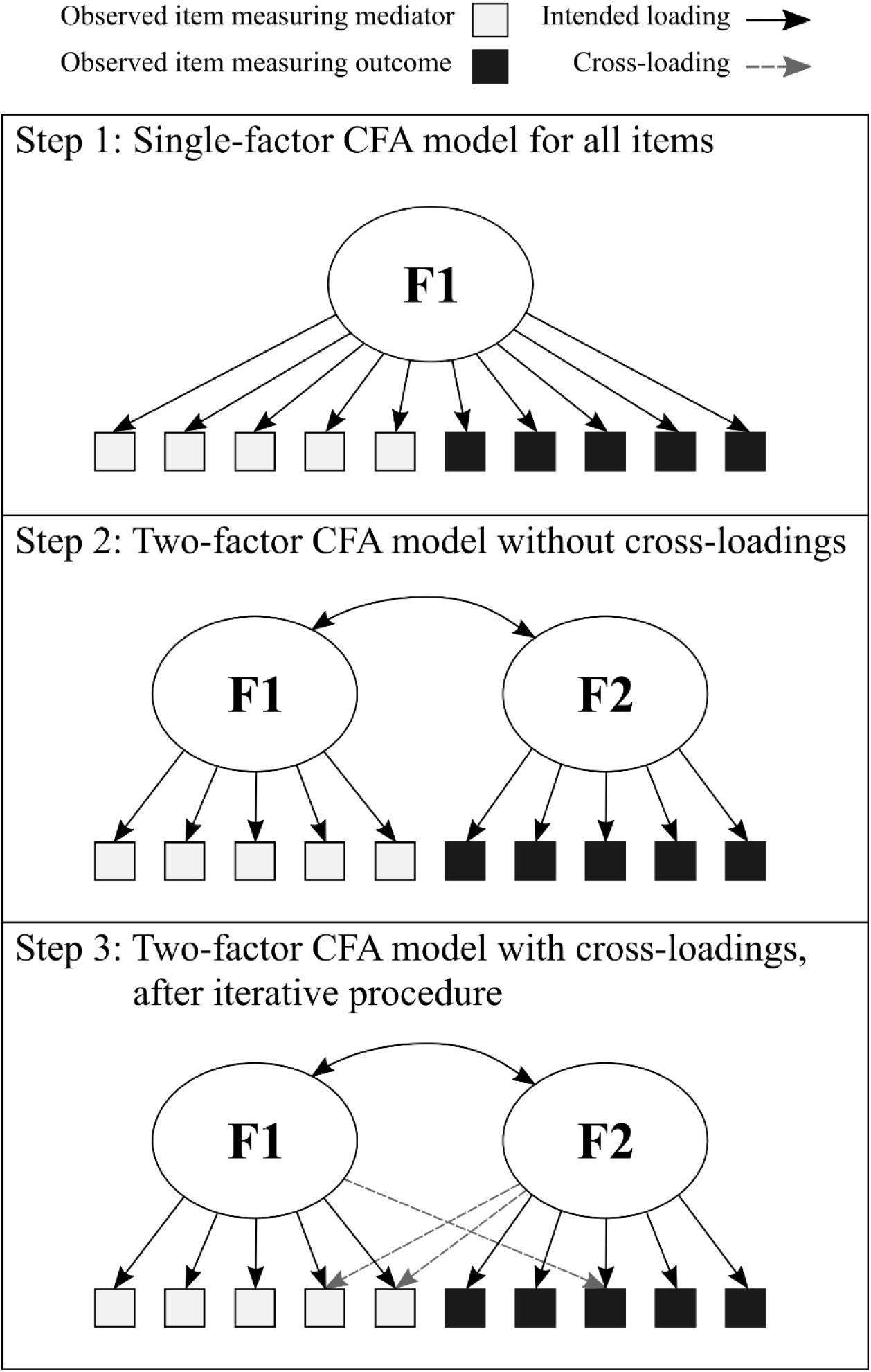
Illustration of model-fitting procedure

Good model fit would have indicated that the observed items for the mediator and outcome could be adequately explained by a single latent variable and that the purported two-factor structure was not required, which was hypothesised to be unlikely.

In Step 2, we estimated a two-factor CFA model, with continuous latent variables for the mediator and outcome measured by their respective observed items (Figure 2, Step 2). Since we expected the mediator and outcome to be related at the structural level, following Chalder et al. (2015), we included a parameter modelling the covariance between each pair of latent variables. Poor fit for the Step 2 model would suggest possible overlap between the mediator and outcome. However, this was tested formally in the next step.

In Step 3, we added cross-loadings to the two-factor model based on the modification indices and improvements in model fit. A cross-loading here refers to a path from the latent variable representing the mediator to an observed item representing the outcome, or vice versa. Overlap was indicated to the extent that the addition of each additional cross-loading item resulted in a statistically significant improvement in overall model fit (tested using the DIFFTEST procedure in Mplus, described above). Starting with the two-factor model from Step 2, above:

i. From the modification indices, select the cross-loading with the highest χ^2^ value and add this to the model.
ii. Use the DIFFTEST procedure to test whether the addition of this cross-loading item produces a statistically significant improvement in overall model fit (p ≤ 0.05).

The above steps were repeated until the additional cross-loading did not result in a statistically significant improvement in model fit, the modification indices did not contain any cross-loadings, or the model failed to converge. For models that failed to converge (which would indicate a poorly-defined model) the previous converging iteration was considered as the final model. In this way, we identified the number of cross-loading items for each mediator-outcome pair (Figure 2, Step 3).

Cross-loading items were classified according to the strength with which the item loaded onto the primary and secondary factors (Table 1). The primary factor was the factor that the item was theoretically intended to load onto, based on prior psychometric analyses; the secondary factor would represent a cross-loading. Factor loadings were classified as strong (0.6 > λ ≤ 1.0), moderate (0.3 > λ ≤ 0.6), weak (0.2 > λ ≤ 0.3) or non-salient (0.0 > λ ≤ 0.2) (Hair et al. 2010). A cross-loading was thus classified as *switched* if at Step 2 it loaded onto the primary factor with a strong or moderate loading, but after Step 3, ‘switched’ to load more strongly onto the secondary factor. An item was *shared* if after Step 3 it loaded with equal strength onto both the primary and secondary factors. A *strong cross-loading* was an item that, after Step 3, continued to load strongly onto the primary factor but also loaded with at least moderate strength onto the secondary factor. Finally, for each mediator-outcome pair we calculated the number of cross-loadings (total, and of each type) and the percentage of cross-loading items (i.e. the total number of cross-loadings after the Step 3, relative to the number of factor loadings at Step 2).

**Table 1:** Classification of cross-loading items, based on factor loading for primary and secondary factors.

| Primary<sup>†</sup> | Secondary<sup>†</sup> | Type for pair |
| --- | --- | --- |
| Non-salient | Non-salient | Non-salient |
|  | Weak | Switched |
|  | Moderate | Switched |
|  | Strong | Switched |
| Weak | Non-salient | No cross-loading |
|  | Weak | Shared |
|  | Moderate | Switched |
|  | Strong | Switched |
| Moderate | Non-salient | No cross-loading |
|  | Weak | No cross-loading |
|  | Moderate | Shared |
|  | Strong | Strong cross-loading (switched) |
| Strong | Non-salient | No cross-loading |
|  | Weak | No cross-loading |
|  | Moderate | Strong cross-loading (switched) |
|  | Strong | Shared |

† Strong = $0.6 > \lambda \leq 1.0$ ; Moderate = $0.3 > \lambda \leq 0.6$ ; Weak = $0.2 > \lambda \leq 0.3$ ; Non-salient = $0.0 > \lambda \leq 0.2$

## Results

In Step 1, almost all single-factor CFA models fitted poorly indicating, as expected, that there were few mediator-outcome combinations where a single latent variable underlying the observed items was consistent with the data. The lowest values of RMSEA and relative χ ^2^ were 0.18 and 21.41, respectively, thus indicating unacceptable fit. One pair had a CFI above 0.95 (for CFQ and the short version of ‘Damage’), however its RMSEA was 0.20. For almost all mediator-outcome pairs CFI was below 0.90 (results available upon request).

Between Steps 2 and 3, cross-loadings were added to most mediator-outcome pairs based on the procedure described above. There were only 4/26 pairs for which no cross-loadings were added (Supplementary Table 2). This was either because the first cross-loading to be tested did not result in a statistically significant improvement in model fit, or because the modification indices contained no cross-loadings. However, most of the cross-loadings added at Step 3 were non-salient. This means they produced a statistically significant improvement in model fit (based on DIFFTEST) but had non-salient factor loadings (below 0.2) onto the secondary factor. Excluding those that were non-salient, only six mediator-outcome pairs exhibited cross-loading, involving a total of 18 cross-loading items. Of these, three were ‘shared’ and 15 were ‘strong’ cross-loadings (Table 2).

**Table 2:** Salient cross-loadings added during the iterative procedure

| Mediator | Outcome<sup>‡</sup> | Number of factor loadings |  | Number of factor loadings added | Type of cross-loading |  |  | Percentage of salient cross-loadings<sup>†</sup> |
| --- | --- | --- | --- | --- | --- | --- | --- | --- |
|  |  | Before iterative procedure | After iterative procedure |  | Non-salient | Shared | Strong cross-loading |  |
| Fear avoidance (short) | CFQ | 14 | 22 | 8 | 4 | 0 | 4 | 29% |
| Avoidance/resting behaviour (short) | CFQ | 14 | 20 | 6 | 2 | 1 | 3 | 29% |
| Damage (short) | CFQ | 14 | 20 | 6 | 2 | 2 | 2 | 29% |
| Fear avoidance | CFQ | 17 | 24 | 7 | 5 | 0 | 2 | 12% |
| Symptom focusing | CFQ | 17 | 23 | 6 | 3 | 0 | 3 | 18% |
| Fear avoidance (short) | PF | 13 | 17 | 4 | 3 | 0 | 1 | 8% |

<sup>†</sup>This is defined as the number of salient cross-loadings relative to the number of factor loadings before carrying out the iterative procedure. <sup>‡</sup> CFQ = Chalder Fatigue Questionnaire; PF = Physical function subscale of the SF-36.

Table 3 presents factor loadings and question wordings for cross-loading items from the six mediator-outcome pairs. In total, there were only 18 instances of cross-loading. Most instances of overlap (17/18) involved items 8 to 11 from the fatigue questionnaire (CFQ). These were questions that assessed whether participants “feel weak” (item 8), “have difficulty concentrating” (item 9), “have problems thinking clearly” (item 10), or “make slips of the tongue when speaking” (item 11). They cross-loaded mostly with the mediators: avoidance/resting behaviour (4 items), damage (4 items), fear avoidance (7 items), and symptom focusing (3 items). The other pair where there was a strong cross-loading was the SF-36 physical function item “Walking several hundred yards” and fear avoidance. Items exhibiting overlap, such as these, indicated that in addition to loading on the latent variable measuring fatigue, combinations of these items also loaded onto the mediator constructs listed above (e.g. avoidance/resting behaviour, damage, and fear avoidance). More precisely, the addition of each cross-loading to the two-factor model resulted in a statistically significant improvement in model fit. The responses to these items, therefore, are influenced by both the outcome and mediator, and thus represent a point of overlap between the two.

**Table 3:** Factor loadings and question wordings for cross-loading items

| Pair |  | Item | Question wording | Factor loadings |  | Type of cross-loading |
| --- | --- | --- | --- | --- | --- | --- |
| Outcome<sup>‡</sup> | Mediator |  |  | Outcome | Mediator |  |
| CFQ | Avoidance/resting behaviour (short version) | CFQ_10 | Do you have problems thinking clearly? | 0.81 | 0.57 | Strong cross-loading |
|  |  | CFQ_11 | Do you make slips of the tongue when speaking? | 0.79 | 0.38 | Strong cross-loading |
|  |  | CFQ_8 | Do you feel weak? | 0.87 | 0.32 | Strong cross-loading |
|  |  | CFQ_9 | Do you have difficulty concentrating? | 0.81 | 0.61 | Shared |
| CFQ | Damage (short version) | CFQ_10 | Do you have problems thinking clearly? | 0.71 | 0.63 | Shared |
|  |  | CFQ_11 | Do you make slips of the tongue when speaking? | 0.73 | 0.41 | Strong cross-loading |
|  |  | CFQ_8 | Do you feel weak? | 0.83 | 0.34 | Strong cross-loading |
|  |  | CFQ_9 | Do you have difficulty concentrating? | 0.71 | 0.61 | Shared |
| CFQ | Fear avoidance | CFQ_10 | Do you have problems thinking clearly? | 0.99 | -0.31 | Strong cross-loading |
|  |  | CFQ_9 | Do you have difficulty concentrating? | 1.00 | -0.34 | Strong cross-loading |
| CFQ | Fear avoidance (short version) | CFQ_10 | Do you have problems thinking clearly? | 0.68 | 0.59 | Strong cross-loading |
|  |  | CFQ_11 | Do you make slips of the tongue when speaking? | 0.70 | 0.39 | Strong cross-loading |
|  |  | CFQ_8 | Do you feel weak? | 0.81 | 0.31 | Strong cross-loading |
|  |  | CFQ_9 | Do you have difficulty concentrating? | 0.69 | 0.59 | Strong cross-loading |
| CFQ | Symptom focusing | CFQ_10 | Do you have problems thinking clearly? | 0.90 | -0.45 | Strong cross-loading |
|  |  | CFQ_11 | Do you make slips of the tongue when speaking? | 0.82 | -0.32 | Strong cross-loading |
|  |  | CFQ_9 | Do you have difficulty concentrating? | 0.91 | -0.43 | Strong cross-loading |
| PF | Fear avoidance (short version) | PF_8 | Walking several hundred yards<sup>†</sup> | 0.83 | -0.31 | Strong cross-loading |

‡ CFQ = Chalder Fatigue Questionnaire; PF = Physical function subscale of the SF-36.
† Does your health limit you in these activities?

## Discussion

In this paper we assessed overlap between latent constructs used to quantify mediators and outcomes in the PACE randomised controlled trial of rehabilitative treatments for chronic fatigue syndrome (White et al., 2011). The purpose of examining overlap was to verify a key assumption of the theoretical mediation model, namely, that these latent constructs were ‘measuring different things.’ Otherwise, rather than evaluating a mediational process, what we would be doing would be more akin to erroneously assessing relationships between repeated measures of our outcome. Adopting a latent variable modelling framework, we assessed whether there were cross-loadings between latent variables representing the mediators and outcomes. Ideally, the observed items for a given mediator should be largely unique and should not simultaneously load onto the outcome. Conversely, the items measuring the outcome should not also serve as indicators of the mediator.

We identified low levels of measurement overlap among the mediators and outcomes in the PACE trial. The single-factor models fitted poorly, suggesting that the items quantifying each mediator-outcome pair could not be explained by a single latent variable. Our analysis suggested many potential cross-loadings, but most of these were non-salient. Out of 26 mediator-outcome pairs, just six indicated cross-loading items, and nearly all of these involved the same four items from a single factor (CFQ). This suggests that the measures used to quantify the mediators and outcomes in PACE were largely distinct.

To our knowledge, no previous studies have examined item overlap in the context of mediation analysis, although several studies have assessed measurement overlap in more general settings. Parker et al. (1991) used exploratory factor analysis (EFA) to assess construct overlap between measures of alexithymia and depression, concluding that “alexithymia was a construct that is distinct and separate from depression” (p. 387). Bagby and Rector (1998) similarly used EFA to assess overlap in personality constructs linked to depression. They found some constructs to overlap (self-criticism and neuroticism) whereas others appeared distinct (dependency and neuroticism). Other studies have used latent variable techniques to assess overlap – e.g. latent class analysis (Mezuk et al., 2013) or item response theory (Hoertel et al., 2015) – but these have adopted differing interpretations of ‘overlap’ and thus are not directly comparable with our approach. For example, Mezuk et al. (2013) defined overlap as the extent to which two sets of observed items measuring depression and frailty identified similar subgroups in their sample. Hoertel et al. (2015) considered whether depressive symptoms (e.g. sadness, fatigue) ‘overlap’ based on differing response profiles in groups of women at different stages of pregnancy. Perhaps closest to our study is the work of Gunzler and Morris (2015) who adopted a latent variable framework to assess overlap between variables measuring depression and variables measuring fatigue and cognitive decline related to multiple sclerosis. Their procedure used MIMIC models (“Multiple Indicator, Multiple Cause”) to test for differential item functioning (DIF), whereas we used confirmatory factor analysis (CFA). Moreover, they took what could be termed a backwards stepwise approach in that they started with a model including all theoretically relevant cross-loadings and *removed* them one-by-one. We started with a model without any cross-loadings and *added* them one-by-one. Both approaches appear valid methods for assessing measurement overlap. Their approach requires that potential cross-loadings are identified a priori based on theory or clinical experience, whereas our approach tests all possible cross-loadings. This exhaustive approach seems preferable when trying to rule out potential overlap, or when lacking strong theoretical expectations as to which precisely items will overlap – as is the case here, given that the chosen constructs were designed to be theoretically distinct.

When assessing mediation it is important to ensure that the mediator and outcome represent distinct constructs. Extensive item overlap would be problematic insofar as it changes the interpretation of the mediation model. In the most extreme case the mediator may no longer represent a distinct third variable. While some studies have sought to reduce overlap by removing similar items (e.g. Segerstrom et al., 2000), this is not always considered or appropriate (e.g. if the psychometric properties of a scale have been validated based on the complete set of items (Bartholomew et al., 2011) then removal of individual items may invalidate the scale).

We found little evidence of mediator-outcome overlap in the PACE trial data, with just 6/26 pairs suggesting overlap, and for each pair no more than 29% of items were affected. It is challenging to define precise thresholds as to what degree of overlap would be considered ‘problematic.’ The proportion of items that exhibit cross-loading is one possible criterion, but the types of cross-loadings also seem important (e.g. the number of ‘shared’ vs. ‘switched’ cross-loadings). It also seems important to consider the theoretical interpretation of the cross-loading items. Are most instances of cross-loading attributable to a single item, or is there widespread cross-loading across many items?

Where substantial overlap is detected – for example, more than half of the items measuring the mediator are simultaneously cross-loading onto the outcome – our interpretation of the mediation analysis should be revised accordingly. In this case, it might be argued that the mediator better represents an earlier measurement of the outcome rather than a distinct construct, so we cannot say with confidence that a *third* variable transmits the effect of our treatment to our outcome. Instead, the situation may be closer to the treatment (T_0_) changing our outcome (T_2_) via a change in an earlier measurement of our outcome (at T_1_). This is an important distinction when it comes to understanding the nature of the processes we are studying – i.e. whether the process is mediational or more akin to repeated measurement of an outcome, which will affect the analysis and interpretation. We recommend that researchers planning mediational analysis using latent traits assess mediator/outcome overlap, and if it is appreciable, either change the measures used, or at least interpret accordingly.

Our analysis used assessments of the mediators at 12 weeks and outcomes at 52 weeks post-randomisation. These time points were chosen to correspond with the measures used in the published mediation analysis (Chalder et al., 2015). An alternative approach, however, might have been to examine construct overlap at baseline (0 weeks). This may be useful when a mediation analysis is planned but follow-up data are not yet available. Another advantage of using baseline measures to assess overlap, in a trial setting, is that they are unaffected by the treatment intervention. However, evidence of a lack of overlap at baseline does not imply absence of overlap at later time points. We suggest then that researchers should either examine overlap based on follow-up assessments that they intend to use for their mediation analysis (once such data become available) or should establish measurement invariance between baseline and follow-up measures. Steps to establish longitudinal measurement invariance are described elsewhere (Fokkema et al., 2013).

### Strengths and limitations

To our knowledge, no studies have addressed the issue of measurement overlap in the context of mediation analysis where the variables of interest are latent (measured via questionnaire). Our study benefited from a large sample size and examined mediators and outcomes that were pre-specified, had been measured on multiple occasions, and had previously been psychometrically validated (Cella et al., 2011; Chalder et al., 1993; McHorney et al., 1993; Ryan et al., 2017). In terms of limitations, the iterative procedure to identify potential cross-loadings made repeated use of the DIFFTEST command in Mplus but no adjustment for multiple comparisons was made. Hoertel et al. (2015), for example, used the Benjamini–Hochberg (1995) procedure to adjust *P*-values produced by DIFFTEST. However, whereas they were attempting to minimize the number of false positives, our analysis was more concerned with false negatives. We wanted to increase the likelihood of detecting cross-loading items, and were thus less concerned by inflated false positive rates due to multiplicity. Also, our approach was somewhat data-driven, in that we used the modification indices to select potential cross-loadings. Importantly, however, the modification indices were not used to select the ‘best’ or ‘correct’ model. The constructs and indicators used to measure mediators and outcomes in PACE were theoretically motivated and purposefully chosen to represent distinct theoretical constructs. We did not use the modification indices to alter these constructs. Rather, the modification indices were used as a computational tool to identify, out of several equally probable cross-loadings, the most influential cross-loading to test first. Ideally, researchers would have a sense about which items will be potentially overlapping. However, in many applications this will not be the case, and the potentially cross-loading items may not be obvious, suggesting the need for a more exploratory approach. This approach is similar to how modification indices are used in the context of a Multiple Indicators Multiple causes (MIMIC) model to identify measurement invariance (e.g. Woods, 2009). An alternative approach might have been to select cross-loadings theoretically, as suggested by Gunzler and Morris. For example, the researcher might select a list of theoretically plausible cross-loadings, and *these* are tested during the iterative procedure, based on their position in the modification indices.

## Conclusions

In conclusion, we found little evidence of construct overlap between mediators and outcomes measured in the PACE trial. This is an important result, supporting the findings of the published mediation analysis (Chalder et al., 2015). Where researchers find more extensive overlap between constructs measuring mediators and outcomes their interpretation of the mediation analysis should be adjusted accordingly, since the mediator can likely no longer be considered a distinct ‘third variable.’ Finally, we reiterate the importance of selecting appropriate and distinct constructs (and questionnaire items) at the point of study design and before data collection. Drawing on relevant theory to inform the selection of distinct constructs will help minimise construct overlap from the outset, allowing studies to obtain more robust conclusions about mediation hypotheses.

## Data Availability

The dataset analysed in the current study are available in the Vivli repository, https://search.vivli.org. Some measures analysed are not publicly available because the authors are still analysing/publishing papers with them.

## List of abbreviations

BDI: Beck Depression Inventory;
CBT: Cognitive behavioural therapy;
CFA: Confirmatory Factor Model;
CFI: Comparative Fit Index;
CFQ: Chalder Fatigue Questionnaire;
DIF: Differential Item Functioning;
EFA: Exploratory Factor Analysis
HAM-D: Hamilton Rating Scale for Depression;
MIMIC: Multiple Indicators Multiple Causes
PF: Physical function;
RMSEA: Root Mean Square Error of Approximation;
RSS: Rumination on Sadness Scale;
SF-36: Medical Outcomes Study Short Form Health Survey;
WLSMV: Robust Weighted Least Squares.

## Declarations

### Ethics approval and consent to participate

This study uses data that have previously been published (Chalder et al. 2015; Ryan et al. 2017). The original PACE study was approved by the West Midlands Multicentre Research Ethics Committee (MREC 02/7/89).

### Consent for publication

Not applicable.

### Competing interests

TC has received royalties from Sheldon Press and Constable and Robinson. Funding for the PACE trial was provided by the Medical Research Council, Department for Health for England, The Scottish Chief Scientist Office, and the Department for Work and Pensions.

The authors declare no other conflicts of interest.

### Funding

This paper represents independent research part funded by the National Institute for Health Research (NIHR) Biomedical Research Centre (BRC) at South London and Maudsley NHS Foundation Trust and King’s College London and Applied Research Collaboration South London (NIHR ARC South London) at King’s College Hospital NHS Foundation Trust. The views expressed are those of the author(s) and not necessarily those of the NHS, the NIHR or the Department of Health and Social Care. EC and SV are fully funded by the NIHR BRC; TC is part-funded by the NIHR BRC; KG is part-funded by the BRC and ARC.

### Authors’ contributions

KG and TC came up with the concept for the study; all authors contributed to the design of the study. EC carried out the data analysis. All authors contributed to the interpretation of results. EC wrote the first draft of the manuscript. All authors contributed to writing the manuscript. All authors gave critical revision of the article and approved of the final version.

## Acknowledgements

The authors acknowledge the other investigators on the PACE trial, Peter D. White and Michael Sharpe, and the help of the PACE Trial Management Group.

---

**Supplementary Table 1.**
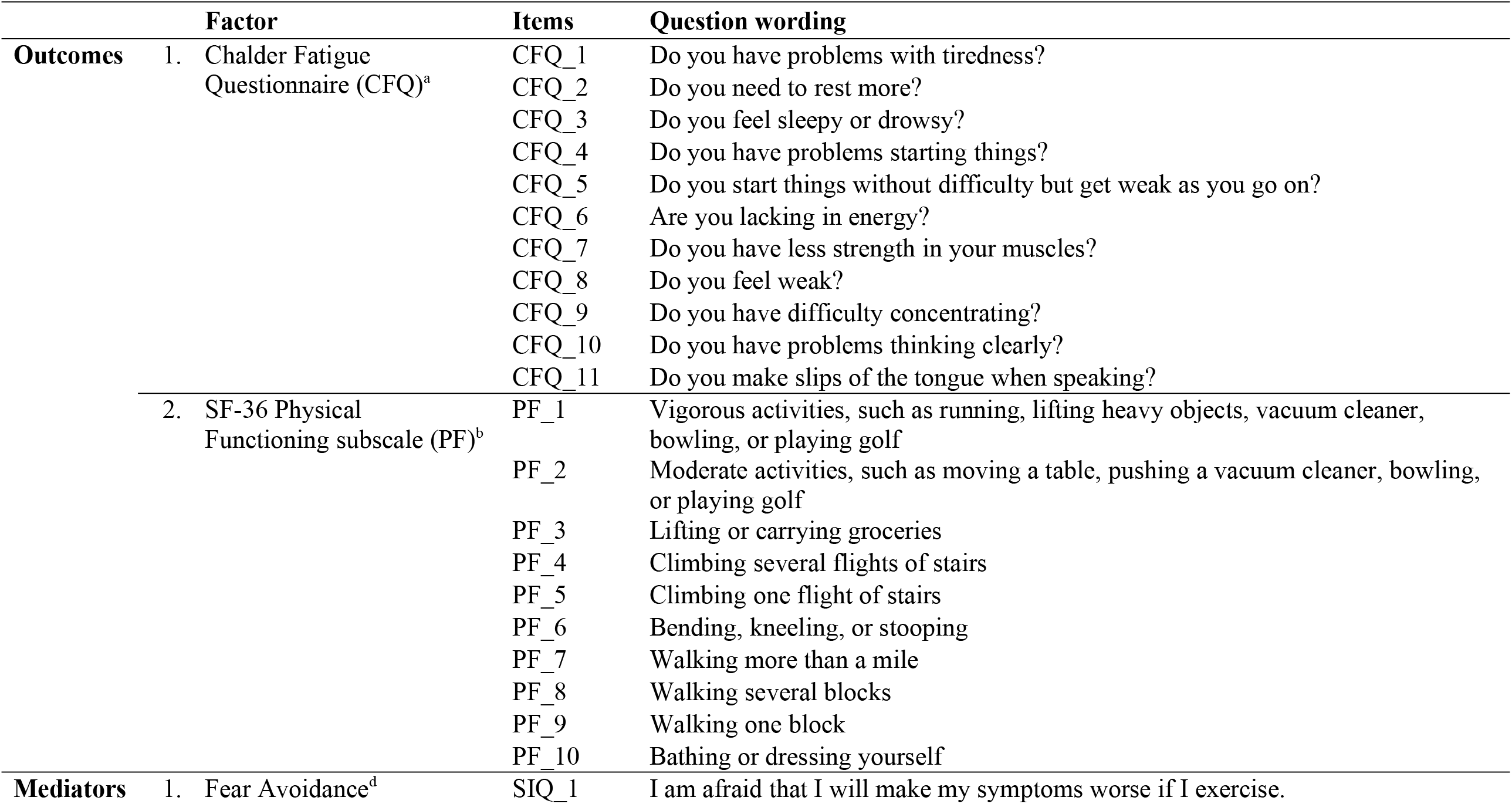
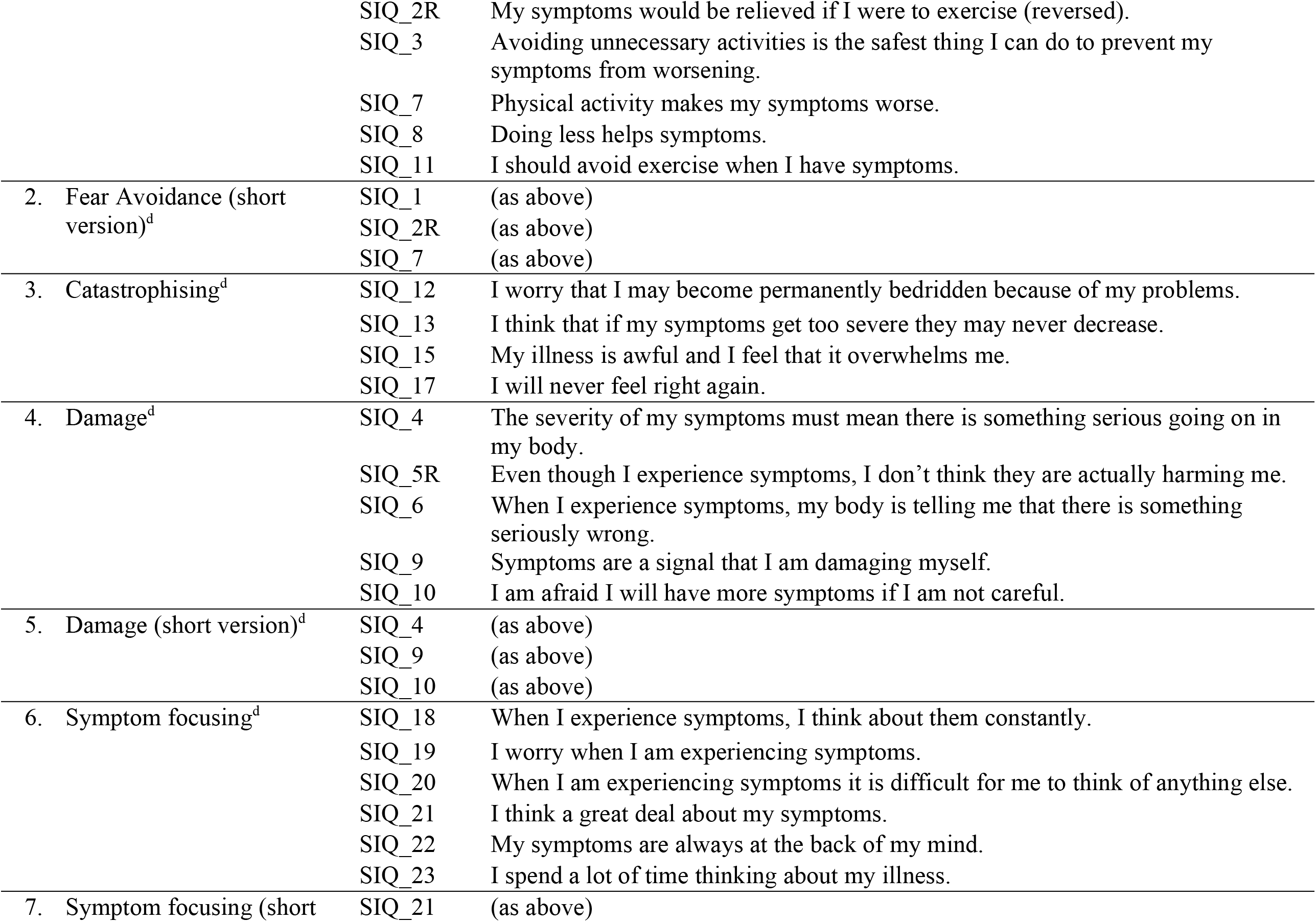
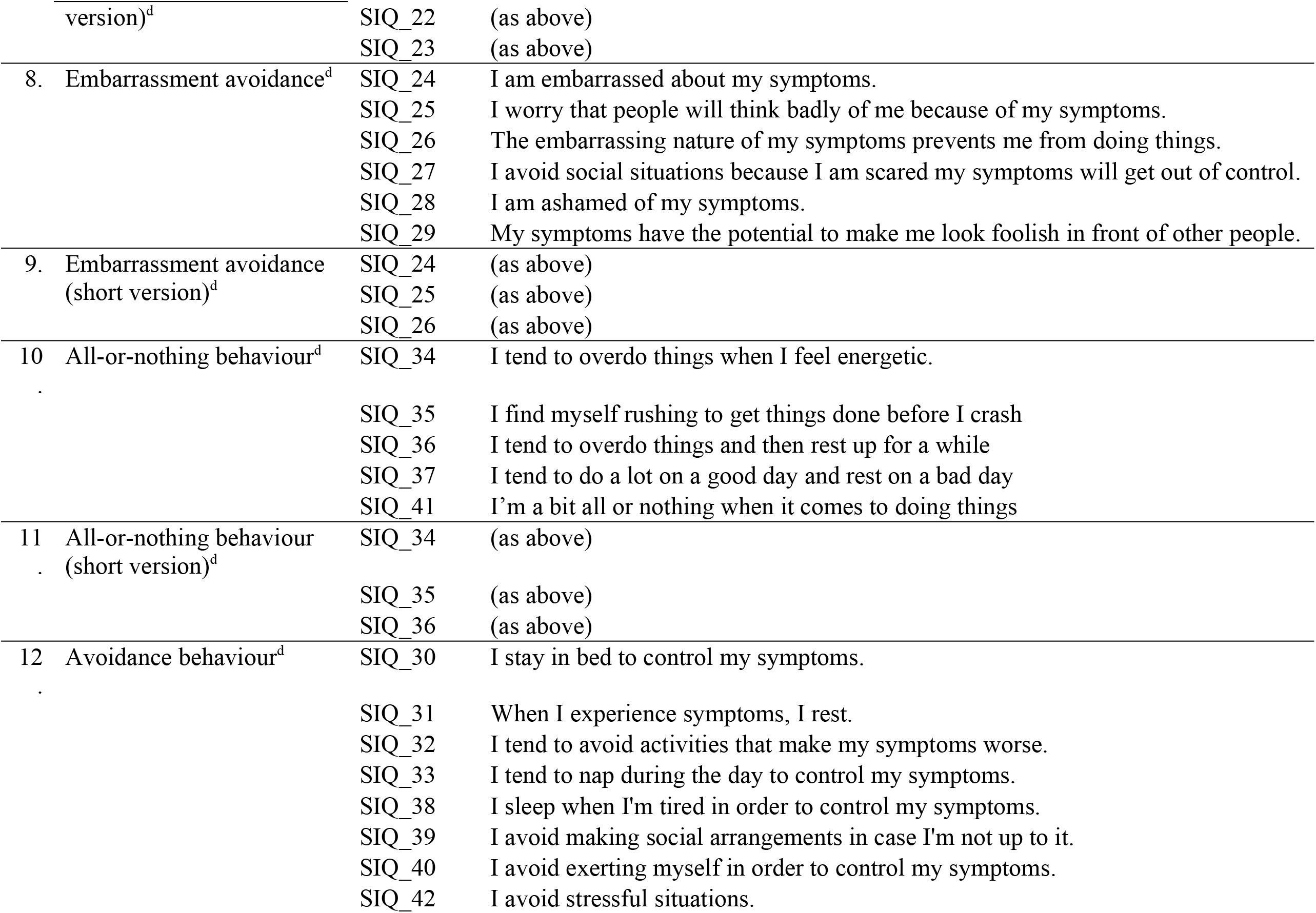
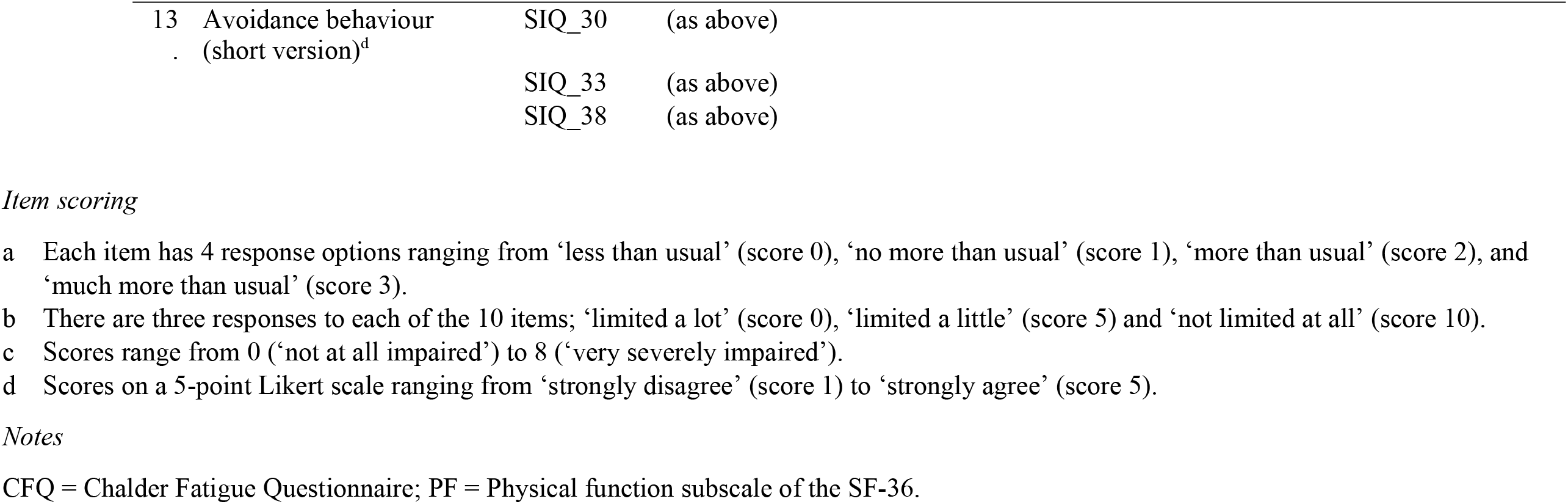
Factor structure and question wordings for PACE mediators and outcomes

**Supplementary Table 2.** All cross-loadings added during the iterative procedure

| Pairing |  | Number of factor loadings |  | Total cross-loadings<br>added by iterative<br>procedure | Type of cross-loadings |  |  |
| --- | --- | --- | --- | --- | --- | --- | --- |
| Mediator | Outcome | Before adding<br>cross-loadings | After adding<br>cross-loadings |  | No cross-loading | Shared | Strong<br>cross-loading |
| Fear avoidance (short) | CFQ | 14 | 22 | 8 | 4 | 0 | 4 |
| Fear avoidance | CFQ | 17 | 24 | 7 | 5 | 0 | 2 |
| Avoidance/resting behaviour (short) | CFQ | 14 | 20 | 6 | 2 | 1 | 3 |
| Damage (short) | CFQ | 14 | 20 | 6 | 2 | 2 | 2 |
| Symptom focusing | CFQ | 17 | 23 | 6 | 3 | 0 | 3 |
| Damage | PF | 15 | 21 | 6 | 6 | 0 | 0 |
| Damage (short) | PF | 13 | 18 | 5 | 5 | 0 | 0 |
| Fear avoidance (short) | PF | 13 | 17 | 4 | 3 | 0 | 1 |
| Fear avoidance | PF | 16 | 20 | 4 | 4 | 0 | 0 |
| Symptom focusing (short) | CFQ | 14 | 17 | 3 | 3 | 0 | 0 |
| Embarrassment avoidance | CFQ | 17 | 20 | 3 | 3 | 0 | 0 |
| All-or-nothing behaviour (short) | PF | 13 | 15 | 2 | 2 | 0 | 0 |
| Avoidance/resting behaviour (short) | PF | 13 | 15 | 2 | 2 | 0 | 0 |
| Symptom focusing (short) | PF | 13 | 15 | 2 | 2 | 0 | 0 |
| All-or-nothing behaviour | PF | 15 | 17 | 2 | 2 | 0 | 0 |
| Catastrophising | CFQ | 15 | 17 | 2 | 2 | 0 | 0 |
| Embarrassment avoidance | PF | 16 | 18 | 2 | 2 | 0 | 0 |
| Catastrophising | PF | 14 | 15 | 1 | 1 | 0 | 0 |
| Embarrassment avoidance (short) | PF | 13 | 14 | 1 | 1 | 0 | 0 |
| Symptom focusing | PF | 16 | 17 | 1 | 1 | 0 | 0 |
| Avoidance/resting behaviour | CFQ | 19 | 20 | 1 | 1 | 0 | 0 |
| Avoidance/resting behaviour | PF | 18 | 19 | 1 | 1 | 0 | 0 |
| All-or-nothing behaviour | CFQ | 16 | 16 | 0 | 0 | 0 | 0 |
| All-or-nothing behaviour (short) | CFQ | 14 | 14 | 0 | 0 | 0 | 0 |
| Damage | CFQ | 16 | 16 | 0 | 0 | 0 | 0 |
| Embarrassment avoidance (short) | CFQ | 14 | 14 | 0 | 0 | 0 | 0 |

CFQ = Chalder Fatigue Questionnaire; PF = Physical function subscale of the SF-36; WSAS = Work and Social Adjustment Scale.

